# Contribution of Beta-Lactamases and Efflux Pumps to Multidrug Resistance in *Pseudomonas aeruginosa* Isolates from ICU Patients in Northeast Brazil

**DOI:** 10.1101/2024.04.23.24306233

**Authors:** Marília S. Maia, Lavouisier F.B. Nogueira, Marco A.F Clementino, Jose Q.S. Filho, Ila F.N. Lima, José K. Sousa, Deiziane V.S. Costa, Jorge L.N. Rodrigues, Luciana V.C. Fragoso, Alexandre Havt, Aldo A.M. Lima

## Abstract

*Pseudomonas aeruginosa* is an opportunistic pathogen with high clinical relevance in intensive care units (ICU) due to its elevated resistance to various antimicrobials, which lead to high morbidity and mortality in patients in critical situations. In this study, we aimed to detect variants of genes encoding β-lactamases and efflux pumps in *P. aeruginosa* isolates resistant to β-lactams, fluoroquinolones and aminoglycosides. All genes belonging to the subfamilies were included in this study: *bla*_SHV_, *bla*_TEM_, *bla*_NDM_, *bla*_KPC_, *bla*_GES_, *bla*_CTX-M_. In addition, we investigated the most relevant variants of the *bla*_OXA_ subfamily and genes belonging to the efflux pumps of the Mex family. We tested 54 isolates of *P. aeruginosa* with a high prevalence of resistance to the antimicrobials piperacillin/tazobactam, ceftazidime, cefepime, imipenem and meropenem. Resistance genes related to carbapenems and spectrum β-lactamases extended were found, which included *bla*_KPC_ genes (81.49%), *bla*_CTXM-2_ (72.22%) and *bla*_CTXM-1_ (66.66%). In relation to the presence of Mex family efflux pumps genes, 100% of positivity were detected. These findings suggest that *P. aeruginosa* isolates exhibit an arsenal of genes encoding β-lactamases able to induce phenotypic patterns of resistance to several antimicrobials commonly used as first-line treatment.

**Author Summary:** Since the introduction of the use of antimicrobials, resistance to antimicrobials has been growing and becoming a global public health problem, as it leads to ineffective treatment and an increased risk of mortality. *P. aeruginosa* is included in the World Health Organization (WHO) critical list of bacteria that have a higher rate of resistance to antimicrobials, requiring constant epidemiological investigation of the strains, especially in hospital environments, to correctly approach them. In this work, we used a methodology that detects 740 variants of different classes of β-lactamases to evaluate the genotype of the study strains against the phenotype found. We evidenced a high prevalence of strains carrying genes related to carbapenems and extended-spectrum β-lactamases, demonstrating a correlation with the phenotypes. Furthermore, we found a 100% positivity rate among the efflux pumps tested belonging to the MEX family.

## Introduction

Antimicrobial-resistant Gram-negative bacteria (GNB) are of particular concern worldwide due to their high morbidity and mortality, particularly in intensive care units (ICUs) [1]. The emergence and spread of microbial resistance are global problems that has profound consequences for public health and the economy [2].

Due to the selective pressure exerted by the frequent use of antimicrobials and the long stay of patients with compromised immune systems, infections related to multidrug-resistant (MDR) microorganisms are prevalent in ICU facilities [3,4]. Healthcare-associated infections (HAIs) caused by MDR are considered a threat to public health, as they limit or even make available therapeutic options unfeasible, which increases the severity and, consequently, the morbidity and mortality rates of those affected, increasing patient length of stay and hospital costs [1].

In this scenario, *Pseudomonas aeruginosa*, an important opportunistic Gram-negative pathogen that causes a variety of infectious diseases, has acquired resistance to several antimicrobials in recent times [5,6]. According to the latest antimicrobial resistance surveillance report from the European Center for Disease Prevention and Control (ECDC), who evaluated *P. aeruginosa* isolates from 2013 to 2016, the average of *P. aeruginosa* isolates with combined resistance (resistance to three or more antimicrobial groups, including carbapenems) was 12.9% [7]. Furthermore, a study carried out in Singapore observed a percentage of 11.5% of health-related infections caused by *P. aeruginosa*, in which 23% of these isolates were resistant to carbapenems [8]. *P. aeruginosa* is included in the “critical” category of the World Health Organization (WHO) priority list of bacterial pathogens for which research and development of new antimicrobials are urgently needed [1].

The main mechanisms involved in antimicrobial resistance in *P. aeruginosa* strains are enzymatic mechanisms, where β-lactamases stand out, and efflux pumps. [5,9]. *P. aeruginosa* isolates contain a variety of β-lactamases, such as the carbapenemases KPC, GES, IMP, VIM, NDM and SPM, and are distributed throughout the world [10,11,12].

Efflux pumps are related to resistance to several antimicrobials, particularly those belonging to the AcrAB-TolC and Mex pump families belonging to the nodulation-division resistance superfamily (RND). Efflux pumps not only mediate intrinsic and acquired microbial resistance, but are also involved in other functions, including bacterial stress response and pathogenicity [13,14,15].

Based on this, we sought to detect variants of genes encoding β-lactamases and efflux pumps, belonging to the RND superfamily, in *P. aeruginosa* isolates resistant to β-lactams, fluoroquinolones and aminoglycosides isolated from patients admitted to an intensive care ward a tertiary care health unit in the city of Fortaleza-CE, with the purpose of tracking the resistance profile of the strains isolated.

## Material and methods

### Obtaining bacterial isolates and identification

For the following study, 259 samples were collected from patients admitted to the clinical ICU of a tertiary care health unit in the city of Fortaleza-CE in the period from April 23, 2019 to May 29, 2021, who presented an infectious condition (defined by the attending ward physicians) involving different anatomical sites. Those microorganisms that were identified as Gram-negative bacteria, resistant to one or more groups of antimicrobials were included: β-lactams, fluoroquinolones, aminoglycosides, macrolides, and glycopeptides. These samples were made available for study upon approval by the research ethics committee of the health unit in question (n° 3.143.258), in which the data were anonymized before accessing them, with the need for consent being waived by the ethics committee in question.

With the aim of diagnosing potential etiological agents, blood culture samples and other noble liquids were inoculated in specific bottles and incubated in the equipment BacT/Alert® 3D (BioMérieux, Marcy l’Etoile, France). The other biological materials were sown in specific culture media using a qualitative or quantitative sowing technique, in order to obtain isolated colonies. Plates were incubated at 37±2°C for 18-24 hours.

The bacterial isolates were identified and tested for their susceptibility to antimicrobials using the automated method VITEK® 2 Compact (BioMérieux, Marcy l’Etoile, France), according to the manufacturer’s recommendations. Minimum inhibitory concentrations were interpreted according to the Clinical and Laboratory Standards Institute [16] and with the Brazilian committee on antimicrobial susceptibility testing [17,18]. For quality control of sensitivity tests, strains from the American Type Culture Collection (ATCC) were used. Specimens that had a resistance profile that fit the research objectives were included in the study.

### Selection of analyzed strains

A total of 258 isolates were analyzed, among which those identified as *P. aeruginosa* were included in this study. Strains of *P. aeruginosa* resistant to one or more groups of β-lactam antimicrobials, fluoroquinolones and aminoglycosides were then used to investigate their genetic resistance profile. MDR was defined as multidrug resistant to ≥1 agent in ≥3 classes of antimicrobials and extending drug resistance (XDR) as drug susceptibility to ≤2 classes of antimicrobials agents.

### Extraction of bacterial DNA

The isolates maintained in trypticase soy broth were incubated overnight in a bacteriological oven at 37°C, at 200 rpm, for subsequent extraction of bacterial DNA. To extract genetic material, the Wizard Genomic DNA Purification extraction and purification kit was used (Promega, Madison, USA), according to the manufacturer’s recommendations.

After extraction, all samples were quantified by spectrophotometry using the NanoDropTM 2000 (Thermo Fisher Scientific, Waltham, Massachusetts, USA) and stored in a -80°C freezer until used in the experiments.

### Detection of resistance-related genes by molecular biology

The selected strains of *P. aeruginosa* were analyzed by real-time PCR (qPCR) using QuantStudio 3® (Applied Biosystems, Massachusetts, USA), using specific primers, for the detection of 13 genes encoding efflux pumps (**Table 1**) and 14 genes encoding β-lactamases described by Nogueira and collaborators [19]. Reactions were standardized using positive controls and negative controls (milliQ water) to determine the most efficient qPCR conditions.

**Table 1.**
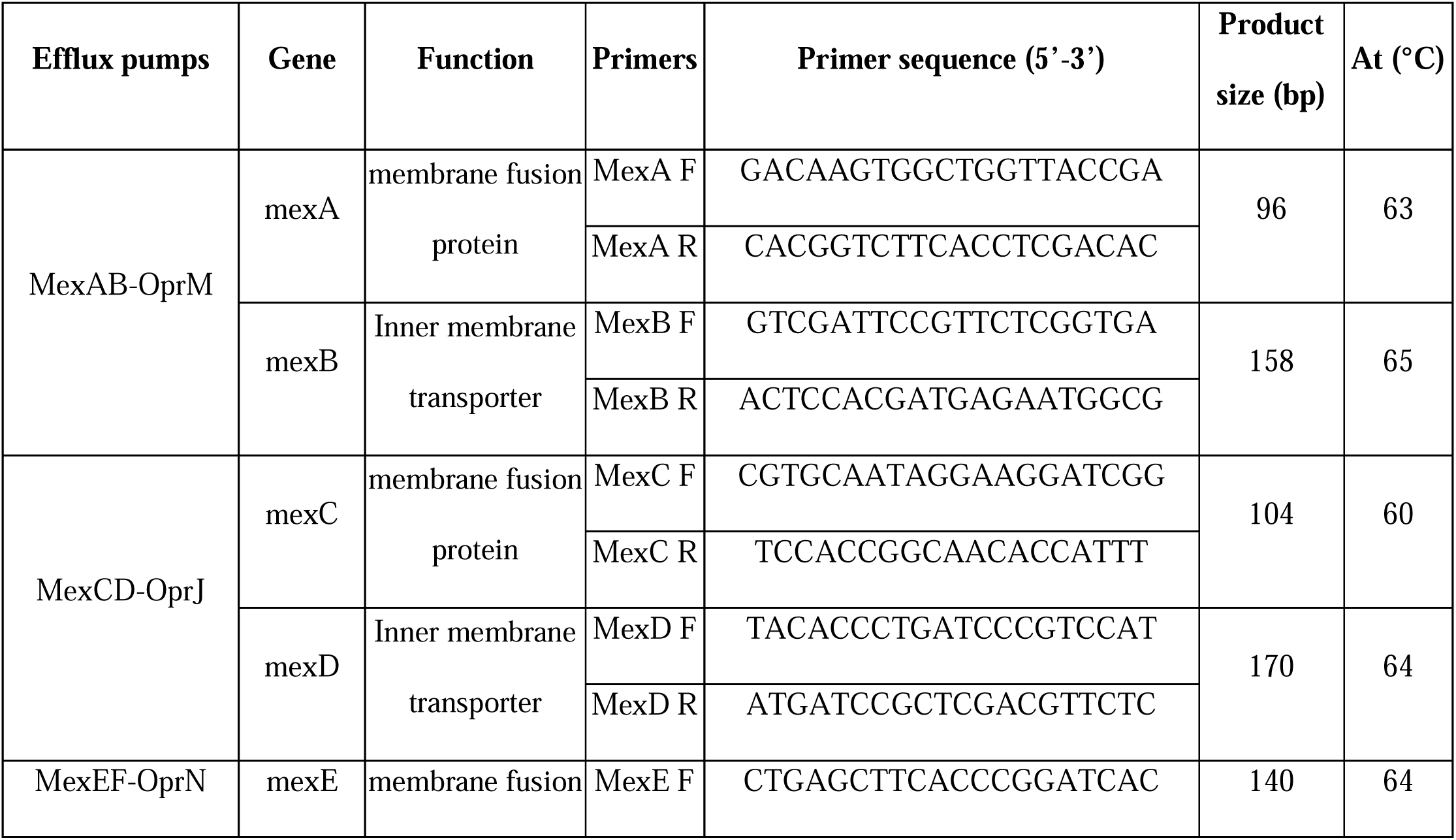
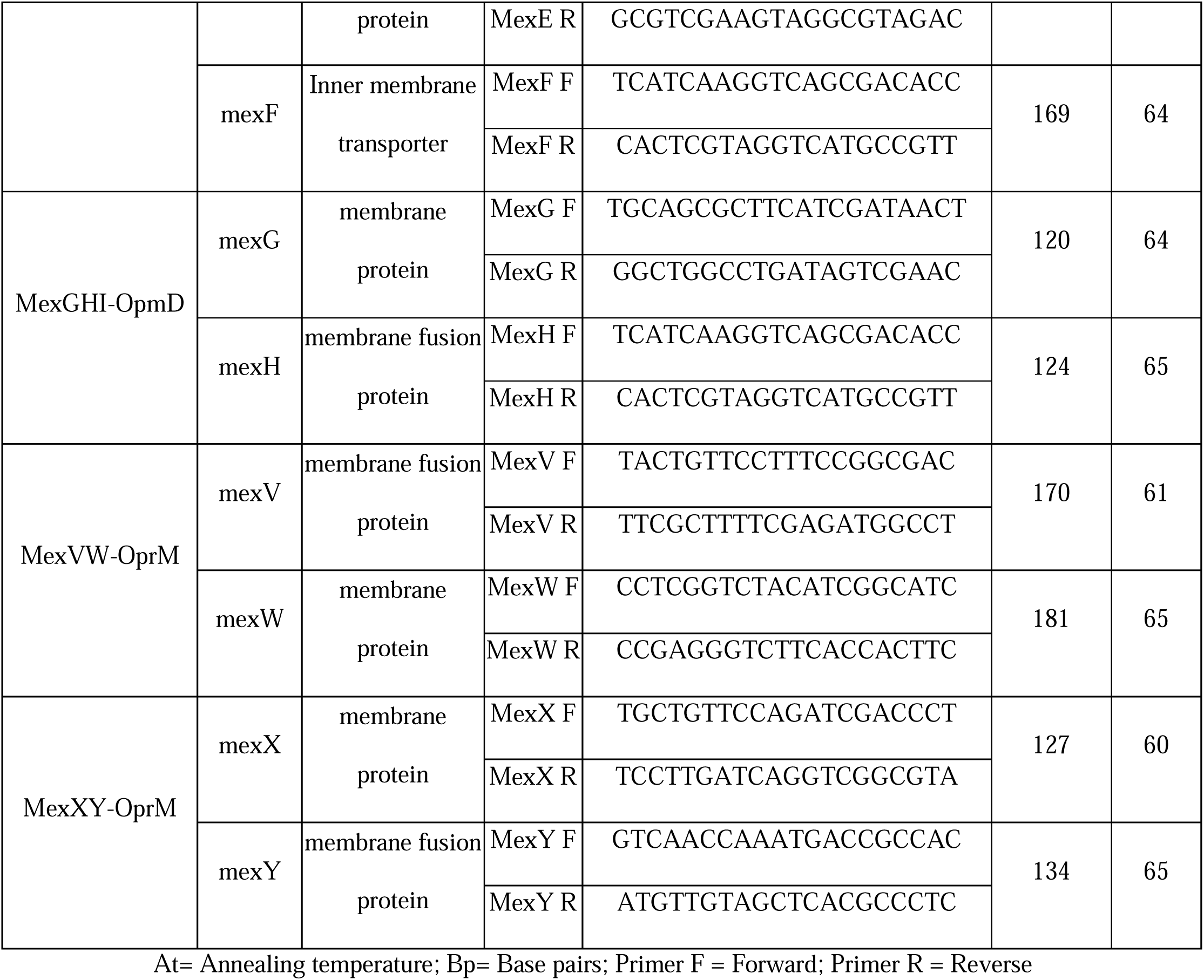
Description of genes related to encoding efflux pumps.

### Data Analysis

Fisher’s test was used to compare the proportion of occurrence of a variable between groups. The confidence interval was 95% and the test was considered statistically significant when P < 0.05, using the IBM Statistical Package for the Social Sciences Version 21 program (SPSS Statistics Software, Nova York, EUA).

## Results

### Prevalence of *P. aeruginosa* in the study

In this study, 258 Gram-negative bacteria were isolated. Among these isolates, (20.9%, 54/258) were identified as *P. aeruginosa*, which were isolated from different anatomical sites of hospitalized patients. Most of the *P. aeruginosa* strains were isolated from tracheal aspirate (74.1%, 40/54), followed by blood samples (11.11%, 6/54), bronchoalveolar lavage (7.40 %, 4/54), abdomen cavity (1.85%, 1/54), tissue fragment (1.85%, 1/54), sacral pressure ulcer (1.85%, 1/54) and tendon (1.85%, 1/54).

### Antimicrobial resistance phenotype tested in *P. aeruginosa* isolates

Fifteen antimicrobials from different classes were tested, in which *P. aeruginosa* isolates obtained a resistance rate of 100% to cefoxitin (32/32), ceftriaxone (25/25), cefuroxime axetil (32/32) and tigecycline (32/32), confirming the intrinsic resistance of this pathogen to these antimicrobials. In addition, most of the *P. aeruginosa* isolates were also resistant to piperacillin/tazobactam (74.4%, 32/43), ceftazidime (63.4%, 33/52) and cefepime (63.4%, 33/52), imipenem (61.2%, 30/ 49), meropenem (56.2%, 27/48), ciprofloxacin (54.9%, 28/51), gentamicin (41.5%, 22/53) and amikacin (28.3%, 15/53) (**Figure 1)**. Interestingly, none of the isolates demonstrate a phenotype of resistance to colistin, ceftazidime/avibactam and ceftolozone/tazobactam.

**Figure 1.**
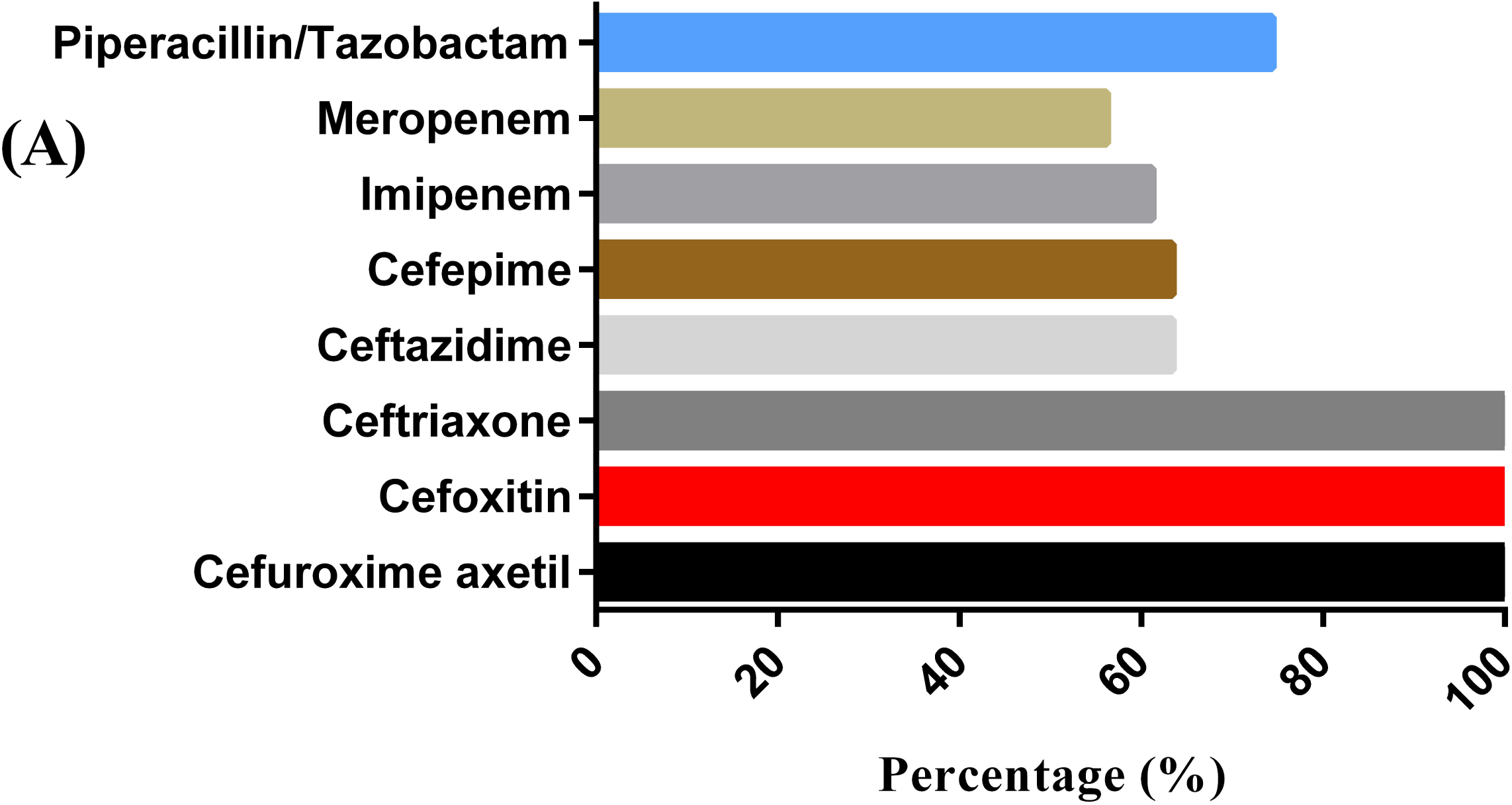
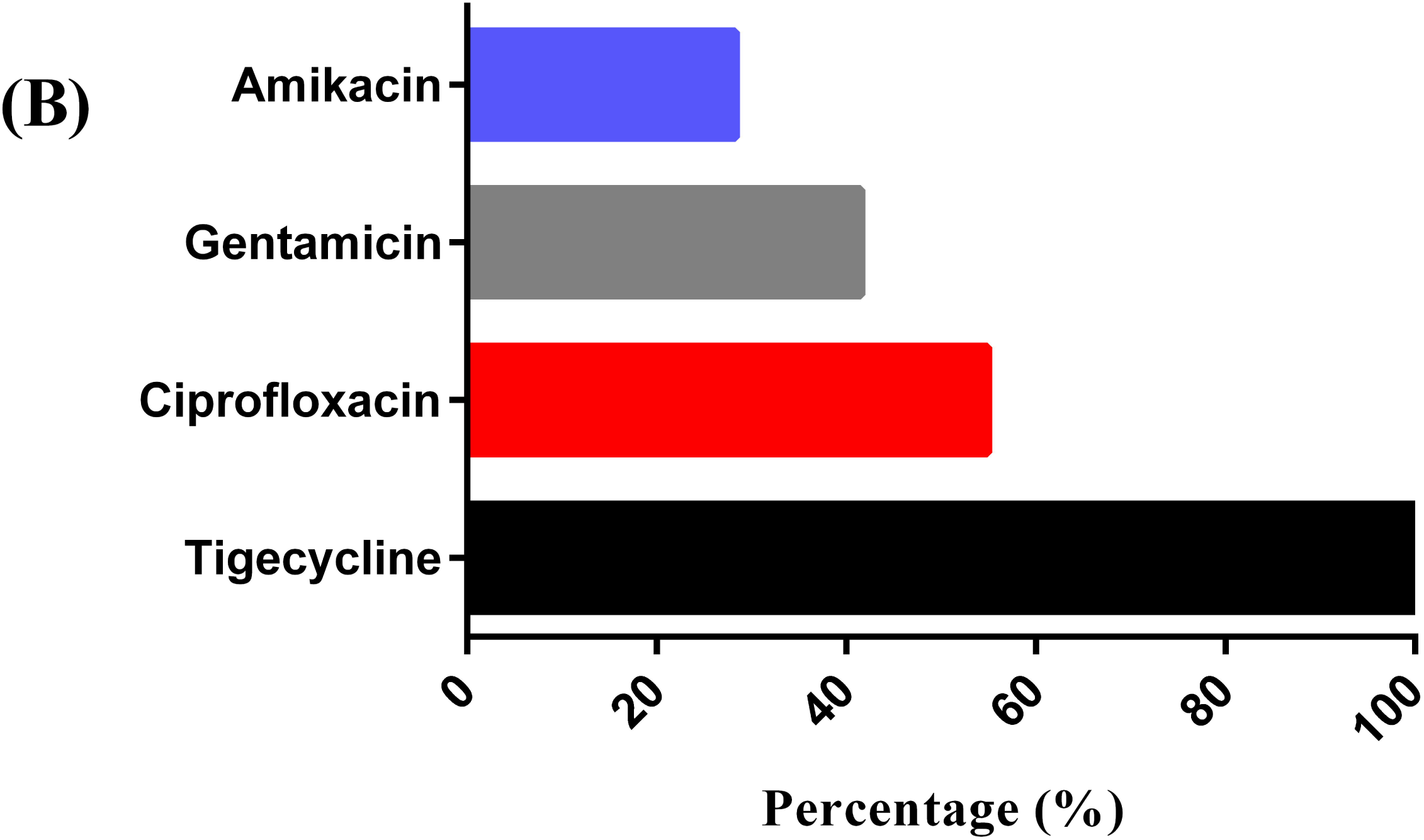
Prevalence of *P. aeruginosa* isolates resistant to β-lactam antimicrobials. (A) and aminoglycosides, quinolones and glycylcycline (B) evaluated, demonstrating a phenotypic profile of resistance to multiple antimicrobials in more than 50% of the isolates, with the exception of colistin, ceftazidime/ avibactam, ceftolozone/ tazobactam, gentamicin and amikacin. The tested isolates of *P. aeruginosa* obtained a percentage of (61.11%, 33/54) for MDR microorganisms, followed by a percentage of (55.5%, 30/54) for XDR microorganisms and a percentage of (46.2%, 25/54) for carbapenem-resistant microorganisms.

### Detection of resistance genes in *P. aeruginosa* isolates by qPCR

For the detection of genes encoding β-lactamases, the highest percentage found was the *bla*_KPC_ gene (81.49%, 44/54), followed by *bla*_CTXM-2_ with (72.22%, 39/54), *bla*_CTXM-1_ (66.66%, 36/54), *bla*_CTXM-5_ (59.25%, 32/54), *bla*_SHV_ (40.74%, 22/54), *bla*_TEM_ and *bla*_OXA-23-like_ (38.89%, 21/54), *bla*_NDM_ (29.6%, 16/54), *bla*_GES_ and *bla*_OXA-51-like_ (24%, 13/54), *bla*_OXA-24/40-like_ (20.4%, 11/54), *bla*_CTXM-3_ (11.11%, 6/54), *bla*_CTXM-4_ (9.26%, 5/54) and there was no detection of the *bla*_OXA-48-like_ gene in the isolates (**Figure 2).**

**Figure 2.**
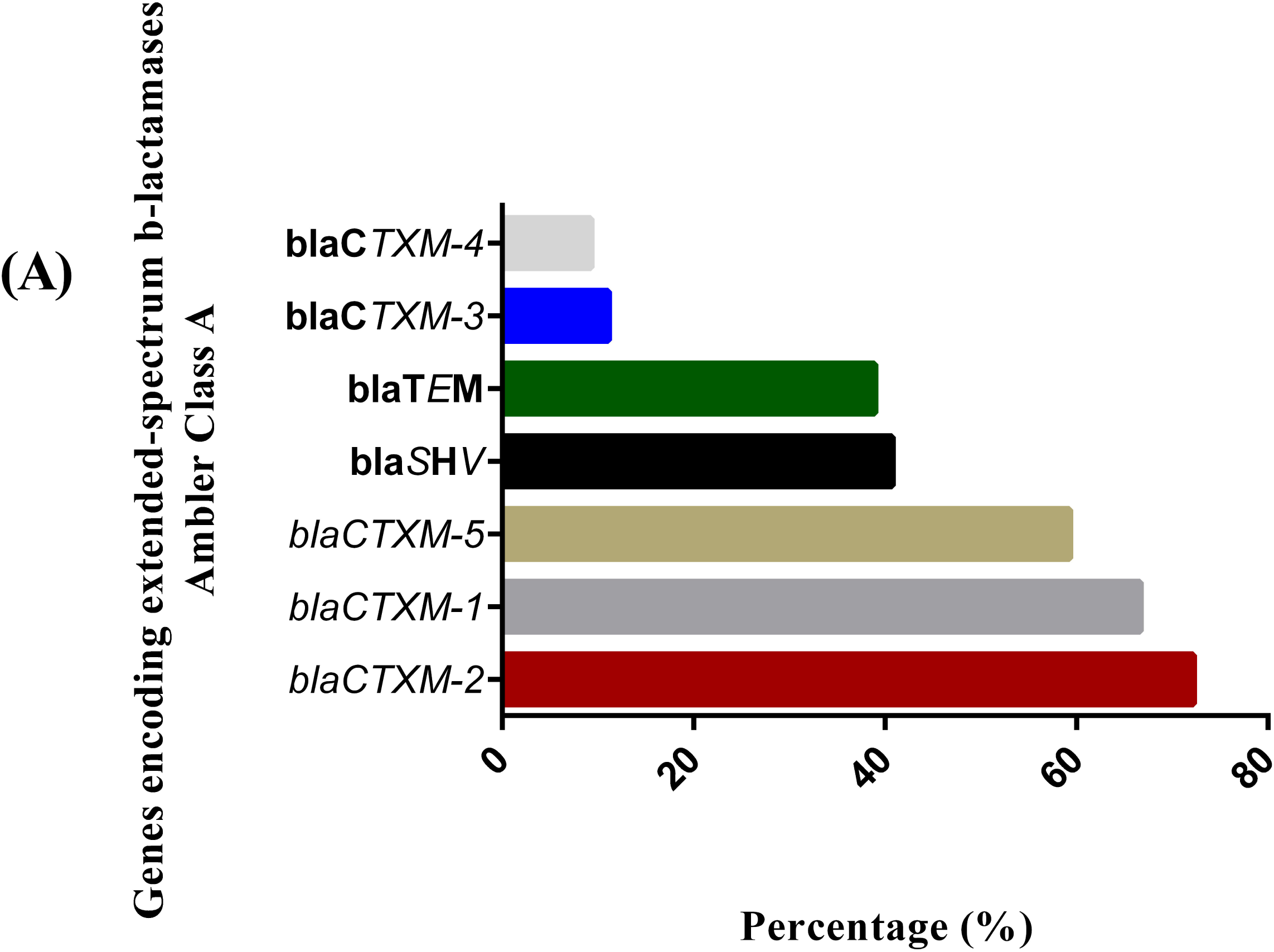
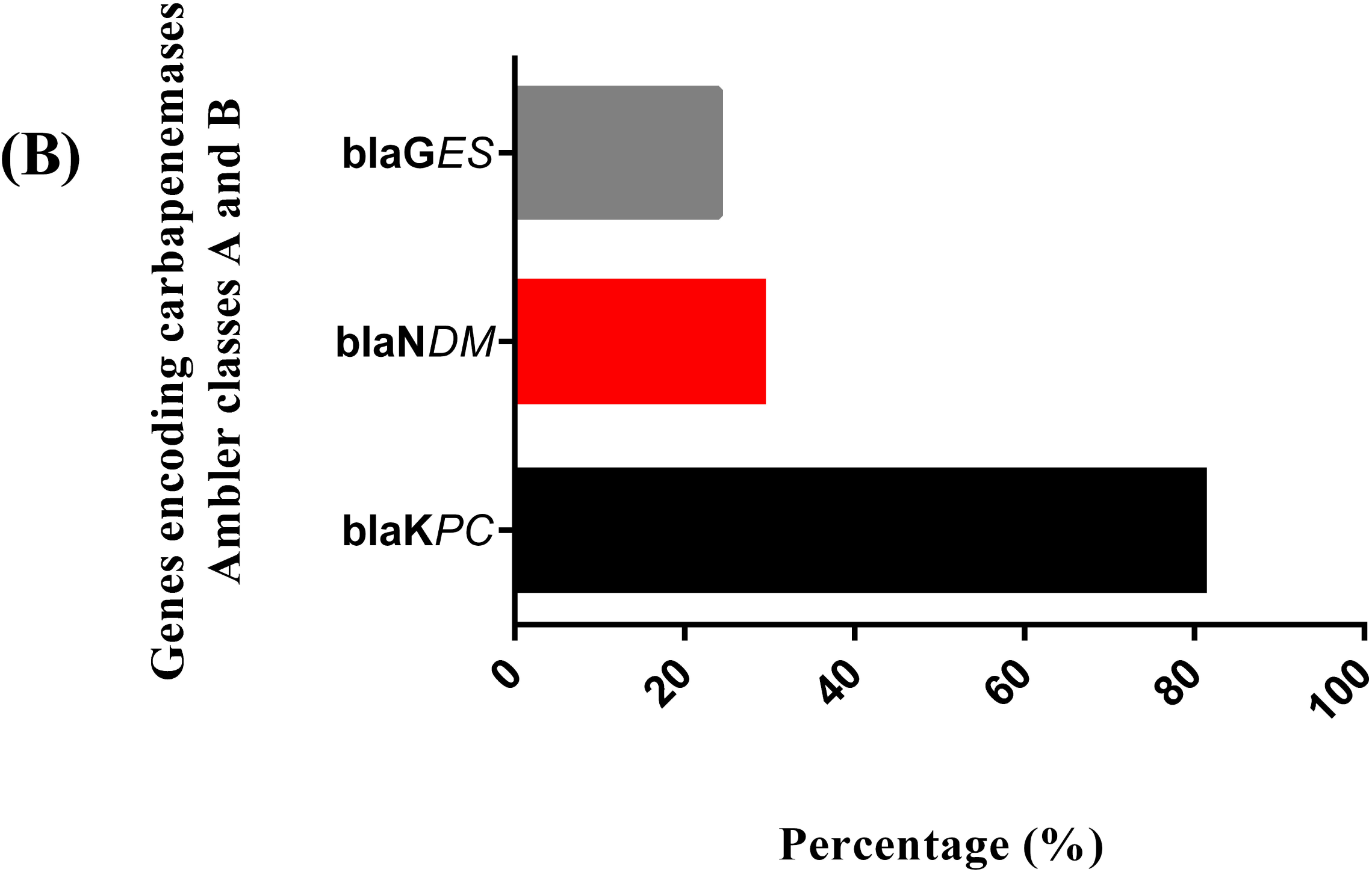
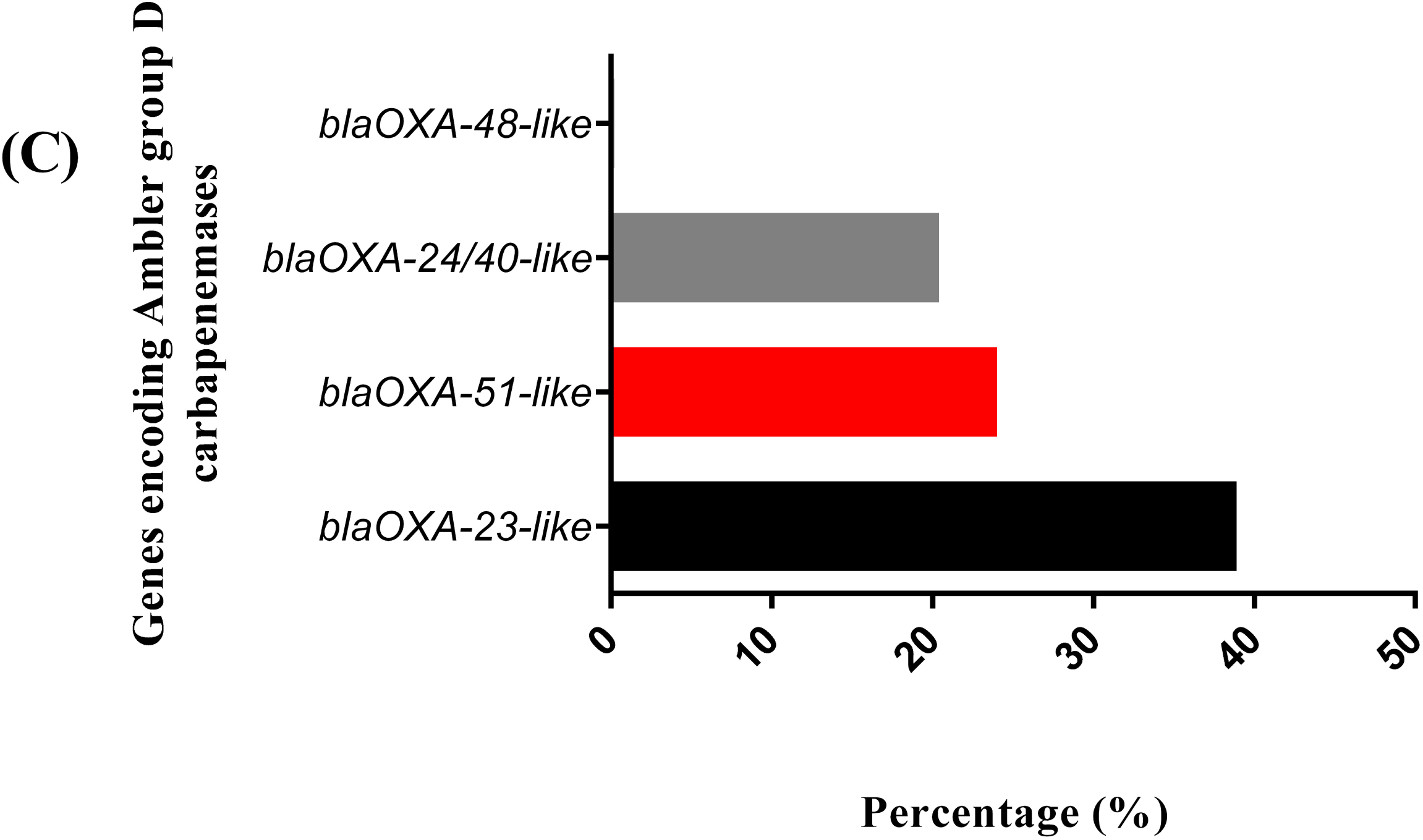
Prevalence of β-lactam resistance genes among *P. aeruginosa* isolates, grouped according to the Ambler classification, (A) in which there is a high detection rate for class A ESBL (*bla*_CTX-M_ do clade 2, 1 and 5 respectively; *bla*_SHV_; *bla*_TEM_), (B) marked presence of the *bla*_KPC_ carbapenemase, (C) in addition to the identification of *bla*_OXA-23-like_, *bla*_OXA-51-like_ and *bla*_OXA-24/40-like_ commonly associated with *Acinetobacter baumannii* strains.

### Association of the presence of resistance genes with the resistance phenotype

The association between the prevalence of the 14 genes encoding β-lactamases and the β-lactam resistance phenotype identified in *P. aeruginosa* isolates was evaluated. The distribution of the percentage of genes detected in the isolates tested for the antimicrobials cefepime and ceftazidime showed a higher positivity rate for the genes *bla*_KPC_ (82.7%, 43/52), *bla*_CTXM-2_ (75%, 39/52), *bla*_CTXM-1_ (69.2%, 36/52), *bla*_CTXM-5_ (61.5%, 32/52) and *bla*_SHV_ (42.3%, 22/52). The presence of the *bla*_TEM_ (P= 0.017; OR 5.66; 95% CI 1.38-23.21) and *blaOXA-23*-like (P=0.041; OR 3.98; 95% CI 1.08 – 14.58) genes were associated with phenotypic resistance to the antimicrobials cefepime and ceftazidime (**Table 2)**.

**Table 2.** Percentage of positivity of genes encoding β-lactamases in *P. aeruginosa* isolates in the groups resistant and susceptible to the antimicrobials cefepime and ceftazidime.

| Coding genes<br>β-lactamases | Total<br>N=52(%) | Resistant<br>N=33(%) | Susceptible<br>N=19(%) | P valor | OR | 95% CI |
| --- | --- | --- | --- | --- | --- | --- |
| *bla*<sub>KPC</sub> | 43 (82.7) | 27 (81.8) | 16 (84.2) | 1.0000 | 0.84 | 0.18 – 3.85 |
| *bla*<sub>CTXM-2</sub> | 39 (75) | 25 (75.8) | 14 (73.7) | 1.0000 | 1.11 | 0.30 – 4.07 |
| *bla*<sub>CTXM-1</sub> | 36 (69.2) | 24 (72.7) | 12 (63.2) | 0.5405 | 1.55 | 0.46 – 5.20 |
| *bla*<sub>CTXM-5</sub> | 32 (61.5) | 19 (57.6) | 13 (68.4) | 0.5579 | 0.62 | 0.19 – 2.05 |
| *bla*<sub>SHV</sub> | 22 (42.3) | 13 (39.4) | 9 (47.37) | 0.3969 | 0.58 | 0.18 – 1.82 |
| *bla*<sub>OXA-23-like</sub> | 21 (40.4) | 17 (51.5) | 4 (21) | **0.041\*** | 3.98 | 1.08 – 14.58 |
| *bla*<sub>TEM</sub> | 20 (38.5) | 17 (51.5) | 3 (15.8) | **0.017\*** | 5.66 | 1.38-23.21 |
| *bla*<sub>NDM</sub> | 16 (30.7) | 10 (30.3) | 6 (31.6) | 1.0000 | 0.94 | 0.27 – 3.19 |
| *bla*<sub>GES</sub> | 13 (25) | 8 (24.2) | 5 (26.3) | 1.0000 | 0.89 | 0.24 – 3.27 |
| *bla*<sub>OXA-51-like</sub> | 13 (25) | 11 (33.3) | 2 (10.5) | 0.0987 | 4.25 | 0.82 – 21.79 |
| *bla*<sub>OXA-24/40-like</sub> | 10 (19.2) | 8 (24.2) | 2 (10.5) | 0.2925 | 2.72 | 0.51 – 14.42 |
| *bla*<sub>CTXM-3</sub> | 6 (11.5) | 3 (9) | 3 (15.8) | 0.6563 | 0.53 | 0.09 – 2.95 |
| *bla*<sub>CTXM-4</sub> | 5 (9.3) | 4 (12.1) | 1 (5.3) | 0.6410 | 2.48 | 0.25 – 24.02 |
| *bla*<sub>OXA-48-like</sub> | 0 | 0 | 0 | 0 | 0 | 0 |

OR= *Odds ratio*; CI= Confidence interval.

The most frequently identified genes in the isolates tested for the antimicrobial piperacillin in combination with the β-lactamase inhibitor tazobactam were *bla*_KPC_, *bla*_CTXM-2_, *bla*_CTXM-1_, *bla*_CTXM-5_, *bla*_SHV_ and *bla*_OXA-23-like_ with percentages of 81.4% (35/43), 74.4% (32/43), 67.4% (29/43), 65.1% (28/43), 41.8% (18/43) and 41.8% (18/43), respectively. There was no statistical significance between the genes studied and the phenotype.

The percentage of positivity of the genes encoding β-lactamases detected in the isolates tested for the antimicrobial imipenem indicated a higher positivity rate for the genes *bla*_KPC_, *bla*_CTXM-2_, *bla*_CTXM-1_, *bla*_CTXM-5_ and *bla*_SHV_ with percentages of 79.6% (39/49), 71.4% (35/49), 67.3% (33/49), 57.1% (28/49) and 38.7% (19/49), respectively. The presence of the *bla*_TEM_ gene achieved statistical significance with phenotypic resistance to the antimicrobial imipenem (P=0.018; OR 5.33; 95% CI 1.28-22.20) (**Table 3)**.

**Table 3.** Percentage of positivity of genes encoding β-lactamases in *P. aeruginosa* isolates in the resistant and susceptible groups to the antimicrobial imipenem.

| Coding genes<br>β-lactamases | Total<br>N=49(%) | Resistant<br>N=30(%) | Susceptible<br>N=19(%) | P valor | OR | 95% CI |
| --- | --- | --- | --- | --- | --- | --- |
| *bla*<sub>KPC</sub> | 39 (79.6) | 24 (80) | 15 (78.9) | 1.0000 | 1.06 | 0.25 – 4.41 |
| *bla*<sub>CTXM-2</sub> | 35 (71.4) | 20 (66.6) | 15 (78.9) | 0.5185 | 0.53 | 0.13 – 2.03 |
| *bla*<sub>CTXM-1</sub> | 33 (67.3) | 19 (63.3) | 14 (73.7) | 0.5412 | 0.61 | 0.17 – 2.18 |
| *bla*<sub>CTXM-5</sub> | 28 (57.1) | 15 (50) | 13 (68.4) | 0.2467 | 0.46 | 0.13 – 1.53 |
| *bla*<sub>SHV</sub> | 19 (38.7) | 10 (33.3) | 9 (47.3) | 0.3774 | 0.55 | 0.17 – 1.80 |
| *bla*<sub>OXA-23-like</sub> | 18 (36.7) | 13 (43.3) | 5 (26.3) | 0.3621 | 2.14 | 0.61 – 7.48 |
| *bla*<sub>TEM</sub> | 18 (36.7) | 15 (50) | 3 (15.7) | **0.018\*** | 5.33 | 1.28 – 22.20 |
| *bla*<sub>NDM</sub> | 15 (30.6) | 7 (23.3) | 8 (42.1) | 1.0000 | 0.87 | 0.27 – 2.81 |
| *bla*<sub>OXA-51-like</sub> | 12 (24.5) | 10 (33.3) | 2 (10.5) | 0.0948 | 4.25 | 0.81 – 22.14 |
| *bla*<sub>GES</sub> | 12 (24.5) | 6 (20) | 6 (31.6) | 0.4975 | 0.54 | 0.14 – 2.02 |
| *bla*<sub>OXA-24/40-like</sub> | 8 (16.3) | 6 (20) | 2 (10.5) | 0.4583 | 2.12 | 0.38 – 11.83 |
| *bla*<sub>CTXM-3</sub> | 5 (10.2) | 3 (10) | 2 (10.5) | 1.0000 | 0.94 | 0.14 – 6.25 |
| *bla*<sub>CTXM-4</sub> | 3 (6.1) | 3 (10) | 0 | 0.2730 | 4.96 | 0.24 – 101.7 |
| *bla*<sub>OXA-48-like</sub> | 0 | 0 | 0 | 0 | 0 | 0 |

OR= Odds ratio; CI= Confidence interval.

The distribution of the percentage of positivity of the genes encoding β-lactamases detected in the isolates tested for the antimicrobial meropenem showed a higher rate for the genes *bla*_KPC_, *bla*_CTXM-2_, *bla*_CTXM-1_, *bla*_CTXM-5_ and *bla*_SHV_ with percentages of 79.6% (39/49), 71.4% (35/49), 67.3% (33/49), 57.1% (28/49) and 38.7% (19/49), respectively. The presence of *bla*_TEM_ (P= 0.034; OR 4.57; 95% CI 1.21-17.23) and *bla*_OXA-51-like_ (P=0.043; OR 5.58; 95% CI 1.06 – 29.20) genes achieved statistical significance with phenotypic resistance to the antimicrobial meropenem (**Table 4)**.

**Table 4.** Percentage of positivity of genes encoding β-lactamases in *P. aeruginosa* isolates in the resistant and susceptible groups to the antimicrobial meropenem.

| Coding genes<br>β-lactamases | Total<br>N=48(%) | Resistant<br>N=27(%) | Susceptible<br>N=21(%) | P valor | OR | 95% CI |
| --- | --- | --- | --- | --- | --- | --- |
| *bla*<sub>KPC</sub> | 38 (79.1) | 21 (77.7) | 17 (80.9) | 1.0000 | 0.82 | 0.19 – 3.40 |
| *bla*<sub>CTXM-2</sub> | 35 (72.9) | 18 (66.6) | 17 (80.9) | 0.3377 | 0.47 | 0.12 – 1.81 |
| *bla*<sub>CTXM-1</sub> | 35 (72.9) | 19 (70.3) | 16 (76.2) | 0.7502 | 0.74 | 0.20 – 2.72 |
| *bla*<sub>CTXM-5</sub> | 28 (58.3) | 13 (48.1) | 15 (71.4) | 0.1437 | 0.37 | 0.11 – 1.24 |
| *bla*<sub>SHV</sub> | 21 (43.7) | 10 (37) | 11 (52.4) | 0.3820 | 0.53 | 0.16 – 1.70 |
| *bla*<sub>OXA-23-like</sub> | 19 (39.5) | 13 (48.1) | 6 (28.6) | 0.2369 | 2.32 | 0.69 – 7.79 |
| *bla*<sub>TEM</sub> | 18 (37.5) | 14 (51.8) | 4 (19) | **0.034\*** | 4.57 | 1.21 – 17.23 |
| *bla*<sub>NDM</sub> | 16 (33.3) | 7 (25.9) | 9 (42.8) | 0.2373 | 0.46 | 0.13 – 1.58 |
| *bla*<sub>OXA-51-like</sub> | 12 (25) | 10 (37) | 2 (9.5) | **0.043\*** | 5.58 | 1.06 – 29.20 |
| *bla*<sub>GES</sub> | 13 (27) | 6 (22.2) | 7 (33.3) | 0.5161 | 0.57 | 0.15 – 2.06 |
| *bla*<sub>OXA-24/40-like</sub> | 8 (16.6) | 6 (22.2) | 2 (9.5) | 0.4371 | 2.71 | 0.48 – 15.11 |
| *bla*<sub>CTXM-3</sub> | 6 (12.5) | 3 (11.1) | 3 (14.3) | 1.0000 | 0.75 | 0.13 – 4.16 |
| *bla*<sub>CTXM-4</sub> | 2 (4.2) | 2 (7.4) | 0 | 0.4973 | 4.21 | 0.19 – 92.73 |
| *bla*<sub>OXA-48-like</sub> | 0 | 0 | 0 | 0 | 0 | 0 |

OR= *Odds ratio*; CI= Confidence interval.

The genes that encode the efflux pumps of the MEX family were evaluated by operon, in which the distribution of the percentage of positivity found in the tested isolates was 100% for the genes MEX-AB, MEX-EF, MEX-GH, MEX-VW, MEX-XY and followed by a percentage of 96.3% for the MEX-CD gene. No statistical tests were performed regarding efflux pumps due to the high positivity of the genes.

## Discussion

*P. aeruginosa* is an opportunistic pathogen with considerable clinical importance in intensive care units, mainly due to its high resistance to several antimicrobials, making effective treatment impossible, leading to high morbidity and mortality in these patients in critical situations [20].

This study found a prevalence of 20.9% of colonization by this microorganism, among Gram-negative isolates resistant to at least one antimicrobial, which represents a similar percentage with other studies, as demonstrated in a report issued by ANVISA [21], where *P. aeruginosa* was the third most isolated microorganism among Gram-negative bacteria reported in bloodstream infections, with a percentage of 16.2%. In the United States, Sader and collaborators, analyzing samples of Gram-negative bacteria isolated from ICU and non-ICU patients, collected in 70 hospital centers, from 2018 to 2020, found a prevalence of 23.5% of *P. aeruginosa* isolated in patients admitted to ICU [22]. Dunphy and collaborators carried out a study in outpatient and hospital environments in regions of Virginia, United States, over a period of one year (2019-2020), collecting several samples, in which they identified a prevalence of 60% of *P. aeruginosa* isolates from different anatomical sites, analyzing 971 isolates collected from 590 patients [23], in which these studies demonstrate a variability in the detection rate of this pathogen.

In this study, the largest number of *P. aeruginosa* isolates was obtained through bronchial lavage secretions with a percentage of 74.1%, similar results were reported in other studies in which *P. aeruginosa* was the most isolated pathogen in lung secretions (bronchial lavage and tracheal aspirate) [23,24]. Respiratory infections caused by this pathogen are quite common, especially in hospital environments, and may be related to the handling of invasive procedures on patients, such as tracheal intubation, aspirations and the need for mechanical pulmonary ventilation [11,20].

In order to identify the phenotypic profile of antimicrobial resistance of *P. aeruginosa* strains isolated in the hospital environment, antimicrobials from different classes were tested, in which the highest resistance rate of *P. aeruginosa* isolates was 100% for cephalosporins of second and third generation cefoxitin, ceftriaxone and cefuroxime axetil. Several studies corroborate this result demonstrating that the only third generation cephalosporin with action on strains of *P. aeruginosa* is ceftazidime [25,26].

In agreement with the findings of this study, research carried out in the United States by Sader and collaborators [22] on *P. aeruginosa* strains collected over a two-year period and a study carried out in Rio de Janeiro over a 20-year period [27], found similar results for the antimicrobials piperacillin with tazobactam, ceftazidime, cefepime and meropenem in strains of *P. aeruginosa*. These studies indicated a growing increase in the rate of resistance to antimicrobials over the years.

Regarding third-generation cephalosporins in combination with β-lactamases inhibitors (ceftazidime with avibactam and ceftolozone with tazobactam), in our study there was no detection of the resistance phenotype. This result is in line with what was expected since ceftolozone with tazobactam is an antipseudomonal antimicrobial that acts on multidrug-resistant strains of *P. aeruginosa* [28,29]. The antimicrobial ceftazidime with avibactam acts on multidrug-resistant *P. aeruginosa* strains that are ESBL, AmpC and KPC positive, as this inhibitor removes the serine residue that is the active portion of the enzyme, however, these antimicrobials do not act on metallo β-lactamases positive strains [30], indicating the action of β-lactamases inhibitors in light of the high positivity of the presence of genes encoding KPC in our study.

The susceptibility rates obtained in our study are in agreement with studies that have reported good activity of ceftazidime associated with avibactam and ceftolozone associated with tazobactam. López-Calleja and collaborators [31] carried out a study in a university hospital located in Spain, in which they analyzed the susceptibility profile of 12 MDR strains of *P. aeruginosa* and 117 XDR strains of *P. aeruginosa* against ceftolozone associated with tazobactam, and detected a susceptibility rate of 92.2% in these isolates. In the same context, research carried out by the Global Antimicrobial Testing Leadership and Surveillance Program, collected 2.521 clinical isolates of *P. aeruginosa* from 41 medical centers in 10 countries in Latin America, from 2017 to 2019, in which they found a rate of 86.9% of isolates susceptible to ceftazidime associated with avibactam [32]. Therefore, these studies corroborate our results, showing that third-generation cephalosporins in combination with β-lactamases inhibitors are a good therapeutic option.

In order to identify the presence and prevalence of genes encoding β-lactamases in the university hospital, we analyzed 14 genes, where the most prevalent gene was *bla*_KPC_ with 81.49% positivity in the isolates. In a study carried out by Hu and collaborators [33], researching strains of carbapenem-resistant *P. aeruginosa* isolated from the intestine, from 2014 to 2019, the presence of the *bla*_KPC-2_ gene was identified in 21.1% of the isolates. Likewise, a prospective and observational cohort study, carried out from 2018 to 2019, analyzed 972 patients admitted with confirmed infection by carbapenem-resistant *P. aeruginosa* in 44 hospitals, distributed across three continents, identifying the presence of the *bla*_KPC-2_ gene in 49% of isolates, demonstrating the circulation of this gene that confers resistance to carbapenems in various parts of the world [34]. Carbapenems have the widest spectrum of action for the treatment of infections caused by multidrug-resistant bacteria, which is why the emergence and spread of carbapenemases has become a major public health problem. Our study detected a rate well above the studies cited, as we used a *bla*_KPC_ primer that detects 19 variants, expanding the detection of this β-lactamase, in contrast to just one variant that the studies above investigated.

To understand the association of the β-lactam resistance phenotype with genes related to β-lactamases, we correlated the phenotype and genotype and found a statistical association between resistance to ceftazidime and cefepime with the presence of the *bla*_TEM_ and *bla*_OXA-23-like_ genes. This can be explained by the fact that the *bla*_TEM_ gene has several variants that are ESBLs, capable of hydrolyzing third and fourth generation cephalosporins [35]. In this case, our study used primers that have the ability to detect 167 variants of the *bla*_TEM_ gene, of which many of these variants are ESBLs. Regarding the *bla*_OXA-23-like_ gene, which encodes an enzyme capable of hydrolyzing all cephalosporins and carbapenems [36,37], was the first-class D carbapenemase to be discovered in a strain of *Acinetobacter baumannii*, in 1985, coincidentally the same year that imipenem was approved for clinical use [38]. Genes belonging to the *bla*_OXA-23-like_ group can be transmitted by plasmids between species, in which studies have detected the role of plasmids and transposons in the dissemination of the *bla*_OXA-23 gene_, which has been found in *Acinetobacter* spp., *P. aeruginosa*, as well as species belonging to the *Enterobacteriaceae* [36,39,40,41].

The presence of the *bla*_TEM_ gene was also associated with resistance to carbapenems, imipenem and meropenem; there is no previous information in the literature that corroborates this result in *P. aeruginosa*. However, a study carried out by Han and colleagues [42], analyzing strains of *A. baumannii* resistant to carbapenems in a university hospital in China, found an association between the presence of the *bla*_TEM_ and *bla*_OXA-23-like_ genes with the carbapenem resistance phenotype. In research carried out in an Iranian hospital, from 2015 to 2017, resistant strains of *P.aeruginosa* were isolated from patients with nosocomal and non-nosocomial infections and the presence of the *bla*_OXA-23-like_ gene was identified in 11.19% of the isolates [43], evidencing the circulation of the OXA group gene in species other than *Acinetobacter*.

Finally, the presence of the *bla*_OXA-51-like_ gene was associated with resistance to meropenem. *bla*_OXA-51-like_ encodes a carbapenemase belonging to Ambler class D, which are enzymes intrinsic to *A. baumannii* and are found naturally in the chromosome of this species [36], however, in 2009, Lee and colleagues reported the presence of the *bla*_OXA-51_ gene in a non-*Acinetobacter* species, in which they identified this gene with an ISAba1 insertion, which was plasmid-encoded and the surrounding sequences suggested that its origin was from *A. baumannii*. In this study, the enzyme encoded by *bla*_OXA-51_ had the capacity to increase the minimum inhibitory concentration (MIC) of meropenem by up to 256 times, thus conferring resistance to it [44]. However, there is only one study in the literature that reports the presence of the *bla*_OXA-51_ gene in *P. aeruginosa* strains, which were isolated from outpatients in Maranhão, Brazil [45]. These studies demonstrate the spread of the *bla*_OXA-51_ gene beyond strains of *A. baumannii*, where this gene was considered a marker for these species.

Regarding the presence of Mex family efflux pumps in *P. aeruginosa* isolates, our study showed a positivity of 96.3% for the gene that encodes the MexCD pump and a positivity of 100% for the rest of the Mex pumps tested. There was no statistical association of efflux pumps in relation to the resistance phenotype due to the high positivity in the isolates, but studies have shown that efflux pumps in *P. aeruginosa* are among one of the main resistance mechanisms that lead to the emergence of MDR strains and XDR. This primary resistance mechanism is responsible for reducing the intracellular concentration of the drug to a subinhibitory concentration at which other resistance mechanisms for specific classes can evolve and be selected, increasing antimicrobial resistance [46,47].

The present study had some limitations, such as: susceptible strains of *P. aeruginosa* were not included in the study for a more detailed epidemiological analysis, no research was carried out into the existence of clones of *P. aeruginosa* circulating in the ICU, no sequencing was carried out positive controls for a more precise validation of all variants that our method aims to detect and the isolates were not tested for all antimicrobials in a homogeneous way due to clinical implications and procedures adopted by the hospital analyzed in the study. However, our work innovated in the use of primers that could detect a greater number of genes encoding β-lactamases, thus expanding their diagnosis.

The *P. aeruginosa* strains isolated in this study showed a high rate of phenotypic resistance to several antimicrobials, in which these strains have a repertoire of genes encoding β-lactamases capable of inducing these phenotypic patterns of resistance to antimicrobials commonly used for these infections, making treatment difficult or even impossible for the patient.

## Author Contributions

Bibliographic review and data collection, Marília S. Maia, Lavouisier F.B. Nogueira, Marco A.F Clementino and Aldo A.M. Lima; Methodology, Lavouisier F.B. Nogueira, Marília S. Maia, Marco A.F Clementino, Alexandre Havt, Ila F.N. Lima, Jorge L.N. Rodrigues, Luciana V. C. Fragoso and Aldo A. M. Lima; Bioinformatics, Lavouisier F.B. Nogueira and Marco A.F Clementino; *In vitro* validation of primers and sample testing, Marília S. Maia, Lavouisier F.B. Nogueira,; Data analysis, Jose Q.S. Filho; Deiziane V.S. Costa, José K. Sousa, and Aldo A.M. Lima.

Written review and editing, Marília S. Maia, Lavouisier F.B. Nogueira, and Aldo A.M. Lima; Supervision, Alexandre Havt and Aldo A.M. Lima; Project administration, Aldo

A.M. Lima; Resource acquisition, Aldo A.M. Lima.

## Data Availability Statement

The data and reports relating to this study will be available via direct request via email to the corresponding author.

